# Impact of Premature Mortality Attributable to Obesity on Years of Life Lost in Brazil

**DOI:** 10.1101/2025.10.16.25338192

**Authors:** Luís Jesuino de Oliveira Andrade, Gabriela Correia Matos de Oliveira, Osmario Jorge de Mattos Salles, Alcina Maria Vinhaes Bittencourt, Luís Matos de Oliveira

## Abstract

**Background:** Obesity represents a substantial global health burden, and obesity-driven premature mortality remains a persistent public health challenge. Although the association between obesity and reduced lifespan is well-established, a precise quantification of the estimated years of life lost (YLL) directly attributable to obesity remains a notable gap in research.

**Objective:** To quantify the societal burden of obesity in Brazil by estimating YLL due to premature mortality between 2014 and 2023.

**Methods:** Life expectancy in Brazil is 76.4 years; thus, premature death was defined as occurring before age 60. This study employed a quantitative epidemiological approach to estimate YLL due to obesity in Brazil (2014–2023). Mortality data were obtained from the Mortality Information System, and demographic projections were sourced from IBGE. YLL was calculated by multiplying deaths by remaining life expectancy (using GBD 2021 reference tables), stratified by age and sex. Statistical analysis was performed using PSPP, and visualizations were generated using Python, with results presented as rates per 100,000 population.

**Results:** The total number of events exhibited an increasing trend over the decade, with a notable peak post-2020, coinciding with the COVID-19 pandemic. The age groups 50–59 and 60–69 consistently showed the highest event frequency, indicating greater prevalence among older populations. Sex-specific analysis revealed variations across age groups and years. While events were rare in younger age categories (1–14), a marked increase was observed from age 15–19 onward. Notably, in older age groups (40–69), females exhibited higher event rates than males, particularly in the 60–69 bracket, suggesting a sex-based disparity in obesity-related mortality.

**Conclusion:** This study highlights an increasing trend in premature mortality due to obesity in Brazil, disproportionately affecting older populations, particularly women. The observed age and sex disparities underscore the need for targeted public health interventions to mitigate obesity-related years of life lost.

## INTRODUCTION

Obesity has emerged as one of the most pressing public health challenges globally, with its prevalence rising dramatically across both high-income and low- and middle-income countries^1^. Characterized by excessive body fat accumulation, obesity is a major contributor to premature mortality through its association with cardiovascular diseases, type 2 diabetes mellitus, certain cancers, and respiratory complications^2^. In Brazil, obesity prevalence has increased substantially over the past three decades, affecting individuals across all age groups and socioeconomic strata^3^. This escalating grievance is compounded by regional disparities in healthcare access, socioeconomic inequalities, and the double burden of malnutrition^4^. Quantifying the societal impact of obesity-related deaths requires robust epidemiological metrics, such as Years of Life Lost (YLL), which estimate the gap between observed age at death and standard life expectancy^5^.

YLL serves as a critical tool for contextualizing premature mortality burden, emphasizing the societal cost beyond traditional mortality statistics^6^. Studies utilizing YLL calculations have revealed up to 40% higher disease impacts than conventional mortality analyses, underscoring the necessity for region-specific assessments^7^. Socioeconomic determinants play a pivotal role in obesity outcomes in Brazil, with income inequality and limited healthcare access contributing to differential mortality rates^8^. Individuals from lower socioeconomic backgrounds face higher risks of obesity-related complications and premature death, attributed to reduced access to healthy foods and limited physical activity opportunities^9^.

Temporal trends reveal concerning patterns, with national surveillance data indicating a 45% increase in obesity-related mortality between 2010 and 2020^10^. The COVID-19 pandemic significantly disrupted obesity management services and exacerbated premature mortality among individuals with obesity^11^. This study seeks to quantify the societal burden of obesity in Brazil by estimating YLL due to premature mortality between 2014 and 2023, providing estimates that account for age, sex, and temporal variations to inform evidence-based policy development aligned with Sustainable Development Goal Target 3.4^12^.

## METHODS

This study utilized a quantitative epidemiological framework to estimate the burden of premature mortality attributable to obesity in Brazil. The core measure employed was the YLL, which provides an estimate of the average years a person would have lived if they had not died prematurely due to obesity-related causes.

### Data Collection

Mortality data were retrieved from the Mortality Information System (SIM) maintained by the Brazilian Ministry of Health, covering the years from 2014 to 2023. The dataset was disaggregated by age, sex, and cause of death, selecting records where obesity was recorded as either the primary or a contributing cause. Demographic information, including population size and age-sex distribution, was obtained from the Brazilian Institute of Geography and Statistics (IBGE) to accurately characterize the population at risk.

### Calculation of Years of Life Lost

YLL was calculated by multiplying the number of deaths within specific age and sex strata by the corresponding remaining life expectancy at the age of death. Life expectancy reference values were adopted from the Global Burden of Disease (GBD) 2021^13^ study conducted by the Institute for Health Metrics and Evaluation (IHME), ensuring standardized and internationally comparable benchmarks. The calculation methodology did not apply age weighting or time discounting, adhering to current ethical considerations and best practices.

### Data Analysis

Data processing and statistical analyses were performed using open-source software PSPP and the Python programming language, with additional calculations performed in Excel. Mortality rates were standardized to per 100,000 population, stratified further by age groups and sex to evaluate trends and disparities throughout the study period.

### Contextual Factors

To complement the quantitative analysis, socioeconomic and healthcare variables such as income distribution, access to health services, and public health policies relevant to obesity prevention and management were incorporated from governmental databases and published literature. This contextualization enabled a more comprehensive understanding of the social determinants influencing premature mortality related to obesity.

### Ethical Considerations

Given the exclusive use of aggregated, anonymized, publicly available data, the study was exempt from requiring individual consent or institutional ethical review, in compliance with Brazilian regulations governing secondary data research.

## RESULTS

### Obesity-Related Premature Mortality in Brazil (2014–2023)

Premature death was operationally defined as occurring before age 60, against a national life expectancy benchmark of 76.4 years. The YLL were estimated by multiplying the number of deaths in each age–sex stratum by the corresponding remaining life expectancy derived from the GBD 2021^13^ reference life tables. Results are reported as age-standardized rates per 100,000 population.

### Temporal Trends in Obesity-Attributable Mortality

A consistent upward trajectory in obesity-related deaths was observed throughout the study period, with a marked acceleration following 2020—coinciding temporally with the onset of the COVID-19 pandemic. Total annual deaths rose from 1,900 in 2014 to a peak of 3,488 in 2021, followed by a modest decline to 2,660 in 2023 (Table 1). This pattern suggests a pandemic-related disruption in chronic disease management and healthcare access, which may have exacerbated underlying obesity-related pathologies.

**Table 1.** Annual distribution of obesity-attributable deaths by age group, Brazil.

|  | AGE RANGE |  |  |  |  |  |  |  |  | TOTAL |
| --- | --- | --- | --- | --- | --- | --- | --- | --- | --- | --- |
| YEAR | 01-04 | 05-09 | 10-14 | 15-19 | 20-29 | 30-39 | 40-49 | 50-59 | 60-69 |  |
| 2014 | 1 | 2 | 3 | 11 | 102 | 287 | 439 | 548 | 507 | 1.900 |
| 2015 | 2 | 1 | 4 | 16 | 89 | 264 | 361 | 579 | 610 | 1.926 |
| 2016 | 0 | 2 | 4 | 12 | 102 | 309 | 489 | 625 | 541 | 2.084 |
| 2017 | 2 | 1 | 2 | 10 | 97 | 270 | 469 | 600 | 603 | 2.054 |
| 2018 | 2 | 3 | 2 | 10 | 101 | 307 | 531 | 616 | 658 | 2.230 |
| 2019 | 1 | 0 | 0 | 10 | 108 | 341 | 550 | 679 | 691 | 2.380 |
| 2020 | 1 | 0 | 1 | 17 | 136 | 447 | 671 | 779 | 838 | 2.890 |
| 2021 | 1 | 1 | 4 | 18 | 160 | 488 | 862 | 964 | 990 | 3.488 |
| 2022 | 2 | 5 | 5 | 23 | 132 | 369 | 617 | 744 | 781 | 2.678 |
| 2023 | 3 | 2 | 5 | 19 | 138 | 367 | 674 | 742 | 710 | 2.660 |

Source: TABNET – Brazilian Ministry of Health

### Age Distribution of Mortality Events

Mortality events were negligible among children and adolescents (ages 1–14), with fewer than five deaths annually across this age range. A progressive increase in event frequency was observed from age 15 onward, with the highest burden concentrated in the 50–59 and 60–69 age groups. These two strata consistently accounted for over 50% of all obesity-attributable deaths each year, underscoring the disproportionate impact of obesity on middle-aged and older adults (Graph 1).

**Graph 1.**
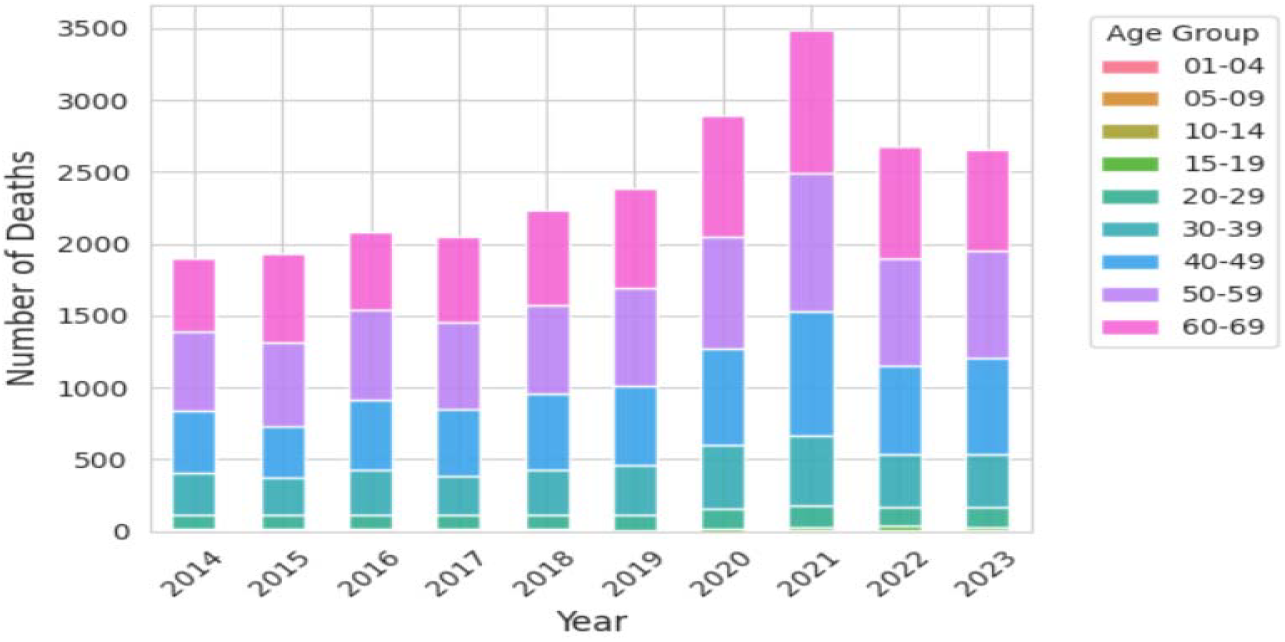
Trends in obesity-related premature mortality by age group, Brazil (2014– 2023). **Source**: Study results.

### Sex-Specific Disparities

Stratified analysis by sex revealed notable gender differences in mortality patterns. While males exhibited slightly higher death counts in younger adult groups (20–49 years), females consistently surpassed males in the 50–69 age brackets (Graph 2). This disparity was most pronounced in the 60–69 group, where female deaths exceeded male deaths in every year of the study period, reaching a difference of 188 additional female deaths in 2021 (Table 2). These findings suggest a sex-based divergence in obesity-related disease progression, healthcare-seeking behavior, or biological susceptibility in later life stages.

**Graph 2.**
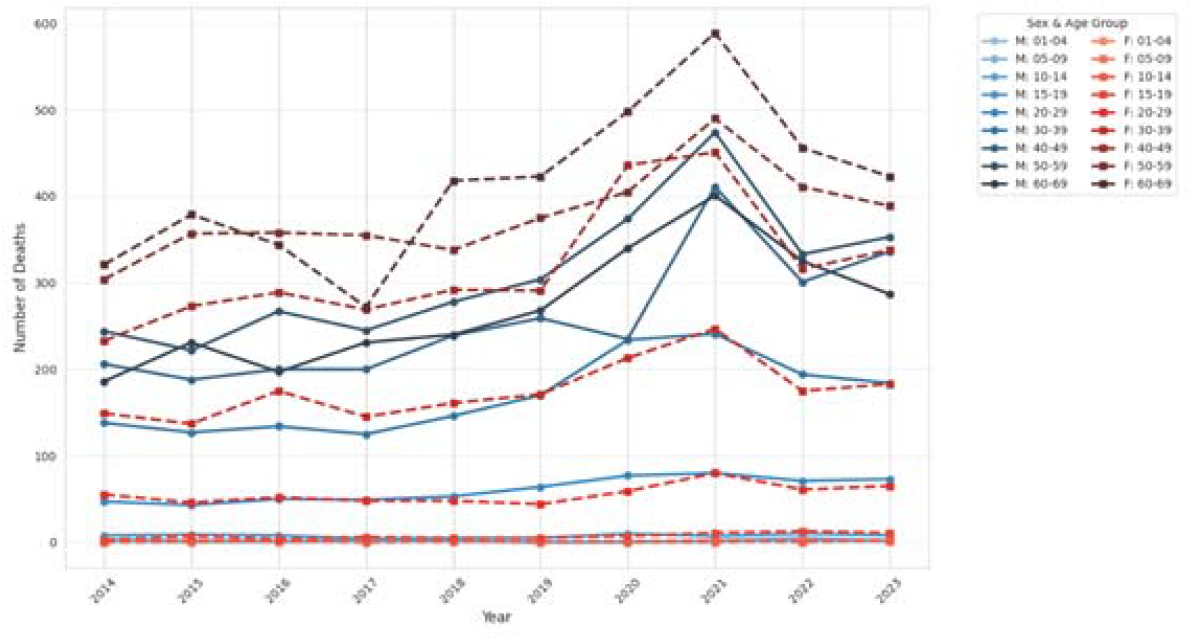
Sex-specific trends in obesity-attributable YLL across age groups **Source**: Study results.

**Table 2.**
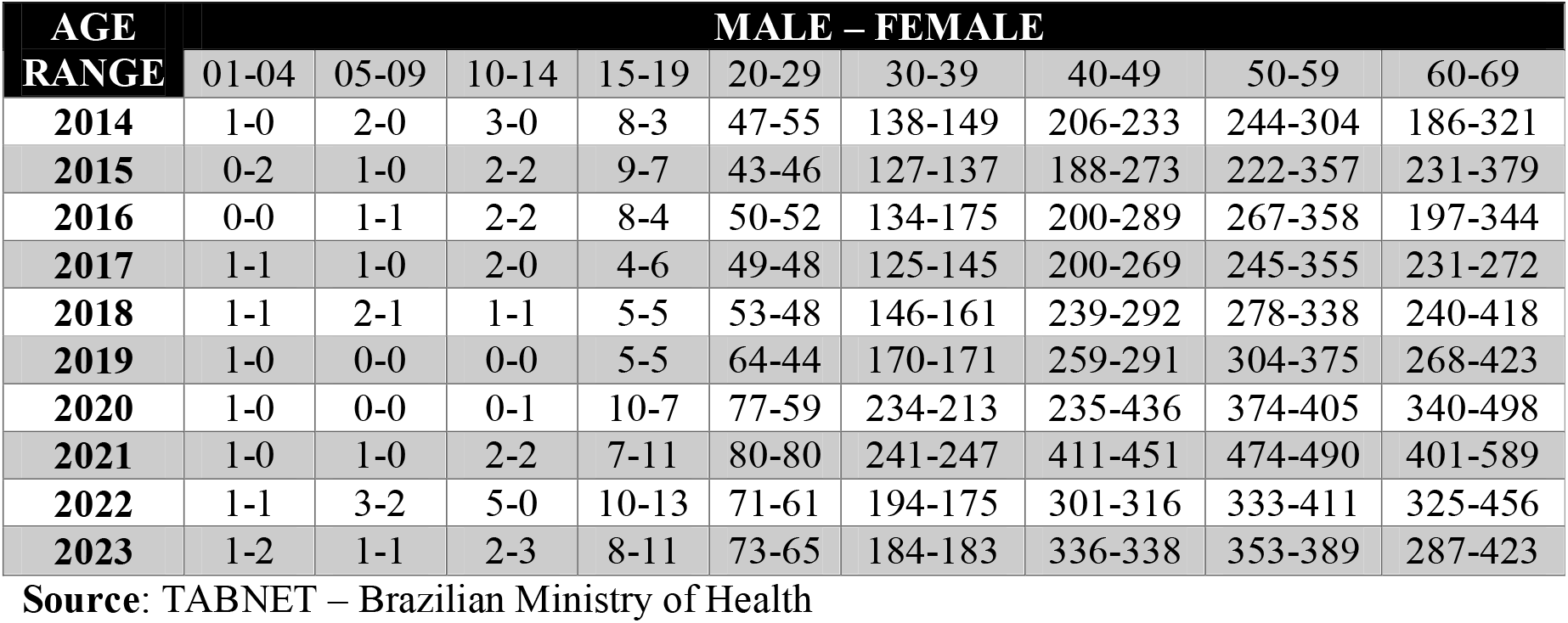
Sex-disaggregated counts of obesity-attributable deaths by age group and year, Brazil, 2014–2023.

Thus, study demonstrates a sustained upward trend in obesity-attributable premature mortality across Brazil from 2014 to 2023, with the highest burden concentrated in middle-aged and older adults. A pronounced gender disparity emerged, as women exhibited consistently higher mortality rates than men in advanced age groups, suggesting sex-specific vulnerabilities in obesity-related health outcomes.

## DISCUSSION

Our study demonstrated an escalation in obesity-driven premature mortality throughout Brazil, disproportionately affecting older demographics and exposing profound sex-based vulnerabilities that demand urgent, evidence-informed public health interventions.

YLL represents a time-based mortality metric that quantifies premature death by calculating the difference between age at death and standard life expectancy at that age, thereby avoiding arbitrary age thresholds while ensuring all deaths contribute proportionally to burden estimates Nearly a decade on - trends, risk factors and policy implications in global obesity^14^. For obesity specifically, YLL calculations reveal marked variations across demographic strata, with younger adults experiencing substantially greater life-year losses than older populations when facing comparable obesity severity levels^15^. Contemporary methodologies, particularly those employed in the Global Burden of Disease Study, calculate YLL by multiplying cause-specific deaths across age-sex-location strata by corresponding standard life expectancies derived from reference life tables^16^. These estimations demonstrate that severe obesity categories produce aggregate burdens exceeding ninety-five million years lost annually, with disproportionate contributions from specific demographic subgroups^17^. Our study aligns with established GBD^13^ protocols, employing standardized life expectancy references while adapting operational thresholds to Brazilian demographic realities, thereby ensuring international comparability alongside contextual epidemiological relevance.

The sustained upward trajectory in obesity-related mortality observed throughout this decade reflects broad epidemiological transitions, wherein non-communicable chronic diseases (NCDs) increasingly overshadow infectious pathologies as the leading causes of death, particularly among aging populations burdened by accumulated metabolic dysfunction^18^. Globally, obesity prevalence has tripled since 1975, with projections indicating that a majority of adults will be overweight or obese by 2050, disproportionately impacting low- and middle-income countries^19^. Our findings align with global trends linking rising obesity mortality to epidemiological shifts toward NCDs, yet reveal a distinct pandemic-associated acceleration, suggesting that disruptions in care during this period intensified pre-existing metabolic vulnerabilities, particularly in populations already facing systemic healthcare inequities. Thus, while worldwide transitions favor chronic metabolic diseases over infectious causes, Brazil’s trajectory demonstrates how health system disruptions compound pre-existing vulnerabilities within resource-constrained settings.

Recent evidence underscores sex-specific vulnerabilities in obesity-related premature mortality, with men consistently exhibiting higher age-standardized YLL rates than women, potentially driven by differential fat distribution, healthcare-seeking behaviors, and hormonal influences^20^. Women experience disproportionate psychopathological burden and obesity-related malignancies, while facing elevated mortality risks despite more frequent treatment-seeking behaviors compared to their male counterparts^21^. Our findings diverge from conventional patterns documented internationally, revealing unexpected female predominance in advanced age groups rather than the typical male excess mortality. This inversion suggests distinctive Brazilian sociocultural factors, healthcare access barriers, or hormonal transitions uniquely amplifying women’s vulnerability.

Our study’s limitations include potential underascertainment of obesity-related deaths due to inconsistent coding practices in death certificates, where obesity is often omitted as a contributing cause. Additionally, defining premature mortality as death before age 60, despite Brazil’s life expectancy of 76.4 years, may underestimate YLL in older adults. Other methodological constraints warrant acknowledgment such as reliance upon secondary mortality data introduces potential misclassification biases, particularly regarding obesity as underlying versus contributing cause of death. Additionally, confounding variables including smoking patterns, socioeconomic stratification, and comorbidity profiles remain unaccounted within aggregated datasets, potentially overestimating direct obesity attribution while obscuring complex causal pathways mediating YLL outcomes.

Thus, our study offers the decade-long quantification of obesity-attributable YLL in Brazil, revealing a concerning upward mortality trend, exacerbated post-2020. Particularly noteworthy is the identification of paradoxical female mortality predominance in advanced age groups, challenging established international patterns and warranting further mechanistic exploration of Brazil-specific sociocultural and healthcare determinants.

## CONCLUSION

This study quantified obesity-attributable premature mortality burden across Brazil, revealing sustained temporal escalation with pandemic-era acceleration. Pronounced age-stratified patterns emerged, concentrating mortality among middle-aged and older demographics. Notably, paradoxical female predominance in advanced age brackets challenges conventional epidemiological paradigms, necessitating targeted interventions addressing sex-specific vulnerabilities to mitigate preventable years of life lost.

## Data Availability

All data produced in the present work are contained in the manuscript

